# Incidence and Severity of Carboplatin-Associated Hearing Loss in Children with Cancer Assessed by the SIOP 2012 Ototoxicity Criteria

**DOI:** 10.64898/2026.05.21.26353442

**Authors:** Aniket Chawla, Sean Carter, Andrew Wood, Sandra Staffieri, Andrew Dodgshun, David D. Eisenstat, Michael Sullivan

**Affiliations:** Children’s Cancer Centre, Royal Children’s Hospital, Parkville, Melbourne, Australia; Murdoch Children’s Research Institute, Parkville, Melbourne, Australia; Department of Paediatrics, University of Auckland, Auckland, New Zealand; Department of Ophthalmology, Royal Children’s Hospital, Parkville, Melbourne; Children’s Haematology/Oncology Centre, Christchurch, New Zealand; Department of Paediatrics, University of Melbourne, Parkville, Melbourne, Australia

**Keywords:** carboplatin, ototoxicity, hearing-loss, platinum, paediatric-oncology, low-grade-glioma, retinoblastoma, neuroblastoma, germ-cell-tumour

## Abstract

**Background:** Platinum-based chemotherapy is known to cause severe and debilitating hearing loss, but unlike cisplatin, the true incidence of carboplatin-induced hearing loss remains unclear. We evaluated functional hearing outcomes in children receiving carboplatin to determine the incidence and severity of ototoxicity.

**Procedure:** We identified a large cohort of children with cancer treated with carboplatin and graded their audiograms using the SIOP ototoxicity scale. Patients with inadequate audiological follow-up, prior hearing loss, or exposure to cisplatin were excluded. Fisher’s exact test, logistic regression, and ROC analyses were performed to investigate associations of demographic, treatment, and exposure-related risk factors with incidence of hearing loss.

**Results:** 200 patients were included, all of whom had been treated with carboplatin. Only nine (4.5%) patients developed clinically significant hearing loss (SIOP grade ≥2). Younger age at first exposure to carboplatin was the only significant predictor of hearing loss (OR = 0.7888, p=0.0241). Age ≤28 months was significantly associated with hearing loss (OR 12.37, p=0.0042). No other risk factors or exposures were statistically significant.

**Conclusions:** Clinically significant carboplatin-associated hearing loss was uncommon (incidence 4.5%). We show that young age is the single-most important risk factor for hearing loss; of nine children who developed hearing loss, eight were aged ≤28 months. Children below this age have twelve-fold higher odds of developing hearing loss compared to those above this age (OR 12.37). These findings will allow physicians to provide more appropriate counselling to families regarding ototoxic risk and support intensified hearing surveillance in young children.

## Introduction

Carboplatin is widely used to treat several childhood cancers, including retinoblastoma, low or intermediate-risk neuroblastoma, germ cell tumours, and low-grade gliomas (LGG). Cisplatin, its structural analogue, is associated with a significant risk of ototoxicity, with documented incidences ranging from 60–90%.^1^ This manifests as a permanent, bilateral, high-frequency sensorineural hearing loss, which may have profound effects on development, communication, cognitive function, and quality of life.^2^

The reported incidence of carboplatin-induced hearing loss varies widely, ranging from 0.9% to 37.5%.^2,3^ (**Table 1**) Younger age at treatment has been associated with a greater risk of hearing loss. Qaddoumi et al. reported that in a cohort of patients with retinoblastoma, children aged <6 months were 21x more likely to develop hearing loss than those aged ≥6 months (p=0.006).^4^ Musial-Bright et al. also noted a significant association between younger age at diagnosis and hearing loss (p=0.016), as well as a positive correlation between cumulative carboplatin dose and hearing loss (p=0.017).^5^ Fetoni et al. reported no relationship between cumulative carboplatin dose and hearing loss.^6^ There has been no dose threshold associated with increased risk of hearing loss in the existing literature. These heterogenous findings likely reflect differences in patient populations, grading scales, surveillance intensity, and concurrent ototoxic exposures.

**Table 1:**
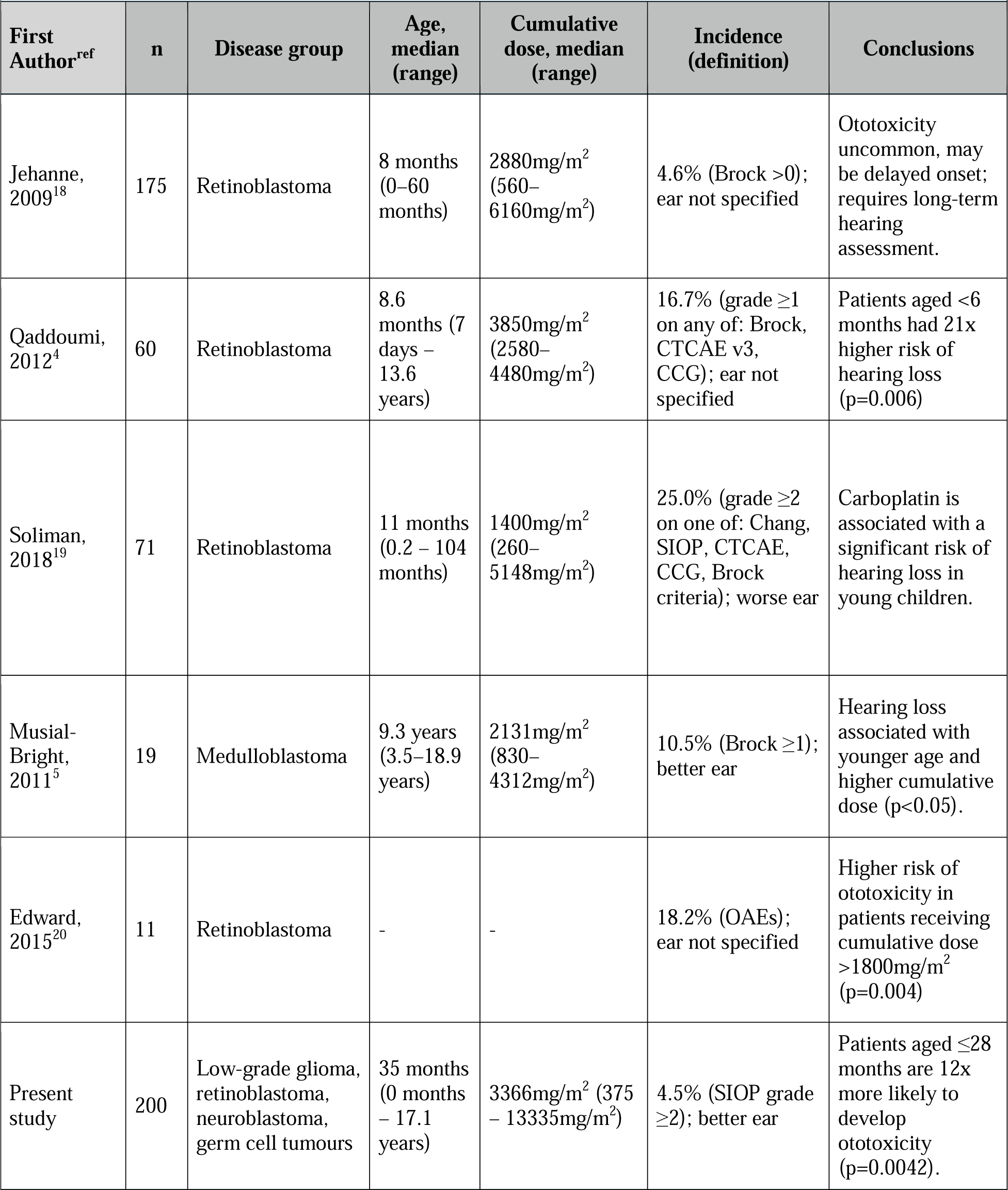
Comparison of Relevant Literature.

| **First Author<sup>ref</sup>** | **n** | **Disease group** | **Age, median (range)** | **Cumulative dose, median (range)** | **Incidence (definition)** | **Conclusions** |
| --- | --- | --- | --- | --- | --- | --- |
| Jehanne, 2009<sup>18</sup> | 175 | Retinoblastoma | 8 months (0–60 months) | 2880mg/m<sup>2</sup> (560–6160mg/m<sup>2</sup>) | 4.6% (Brock >0); ear not specified | Ototoxicity uncommon, may be delayed onset; requires long-term hearing assessment. |
| Qaddoumi, 2012<sup>4</sup> | 60 | Retinoblastoma | 8.6 months (7 days – 13.6 years) | 3850mg/m<sup>2</sup> (2580–4480mg/m<sup>2</sup>) | 16.7% (grade ≥1 on any of: Brock, CTCAE v3, CCG); ear not specified | Patients aged <6 months had 21x higher risk of hearing loss (p=0.006) |
| Soliman, 2018<sup>19</sup> | 71 | Retinoblastoma | 11 months (0.2 – 104 months) | 1400mg/m<sup>2</sup> (260–5148mg/m<sup>2</sup>) | 25.0% (grade ≥2 on one of: Chang, SIOP, CTCAE, CCG, Brock criteria); worse ear | Carboplatin is associated with a significant risk of hearing loss in young children. |
| Musial-Bright, 2011<sup>5</sup> | 19 | Medulloblastoma | 9.3 years (3.5–18.9 years) | 2131mg/m<sup>2</sup> (830–4312mg/m<sup>2</sup>) | 10.5% (Brock ≥1); better ear | Hearing loss associated with younger age and higher cumulative dose (p<0.05). |
| Edward, 2015<sup>20</sup> | 11 | Retinoblastoma | - | - | 18.2% (OAEs); ear not specified | Higher risk of ototoxicity in patients receiving cumulative dose >1800mg/m<sup>2</sup> (p=0.004) |
| Present study | 200 | Low-grade glioma, retinoblastoma, neuroblastoma, germ cell tumours | 35 months (0 months – 17.1 years) | 3366mg/m<sup>2</sup> (375 – 13335mg/m<sup>2</sup>) | 4.5% (SIOP grade ≥2); better ear | Patients aged ≤28 months are 12x more likely to develop ototoxicity (p=0.0042). |

Concurrent ototoxic exposures complicate assessment of carboplatin’s true ototoxic potential. Cranial radiotherapy contributes to hearing loss due to its impact on the cochlea, with a 5% risk of hearing loss with cochlear radiotherapy doses ≤35Gy, and a 30% risk at 50Gy.^7^ Vincristine has also been recently identified in three studies as compounding cisplatin-induced hearing loss.^8–10^ A 2021 study of 1481 patients that showed cisplatin-induced ototoxicity was 3.55x more likely in patients receiving vincristine (OR 3.55, p<0.0001), and a 2023 study demonstrated that co-administration of vincristine increased platinum-induced ototoxicity from 22% to 49% (p<0.001).^8,9^ This may be due to its neurotoxic impact on the medial olivocochlear system and outer hair cells. Cerebrospinal fluid (CSF) shunting has also been identified as a significant risk factor for platinum-induced hearing loss (p=0.0008).^11^ Lastly, aminoglycosides and loop diuretics are widely-recognised contributors to ototoxicity.^12–13^ Patients receiving carboplatin often simultaneously receive a combination of these interventions.

The SIOP ototoxicity scale is the current internationally agreed-on scale for grading platinum-induced hearing loss.^14^ This grading system uses absolute hearing thresholds rather than changes from baseline, recognising that baseline audiology is not always attainable in ill children, and has demonstrated greater sensitivity than the previously common Brock criteria.^15^ Its use in this study provides a contemporary evaluation of carboplatin-associated hearing loss across a range of paediatric malignancies. Relevant risk factors investigated include age, cumulative dose, and other ototoxic treatments, such as vincristine, cranial radiotherapy, CSF shunting, aminoglycosides, and diuretics. Our findings will enable clinicians to accurately discuss the risk of hearing loss with patients and families and arrange necessary audiological follow-up.

## Methods

Ethical approval was obtained from the Royal Children’s Hospital, Research Ethics & Governance office, prior to commencement of data collection (REG 3935).

We conducted a retrospective review of all patients aged 0 –18 years diagnosed with LGG, retinoblastoma, germ cell tumours, or low/intermediate-risk neuroblastoma, at the Royal Children’s Hospital, Melbourne, from 1995 –2024. Patients who received carboplatin and post-exposure audiological assessment were included. Patients without pre-exposure (baseline) audiological assessments were also included, provided there were no documented clinical concerns regarding their auditory function. Exclusion criteria included cisplatin exposure, unavailable audiology reports, abnormal hearing at baseline, and uninterpretable post-exposure audiology results.

Data was collected on patients’ age, sex at birth, disease type, age at diagnosis, and chemotherapy exposure, including dates of administration of carboplatin and vincristine, corresponding doses, and Body Surface Area (BSA) at time of administration. Cumulative doses were calculated based on BSA for all patients regardless of age, to allow consistent comparison across the cohort. Exposure to cranial radiotherapy, CSF shunts, aminoglycosides, and loop diuretics by the time of last audiogram was recorded. All audiograms performed to date were individually reviewed by one investigator (AC) and assessed against the SIOP ototoxicity criteria. Each ear was individually graded, and the better ear was used for data analysis to reflect functional hearing. Clinically significant hearing loss was defined as SIOP grade ≥2, as grade 1 reflects milder high-frequency thresholds that may not cause functional hearing loss in children; grades 0-1 were defined as “normal hearing” and grades ≥2 as “hearing loss”. Grades at the last audiogram were used to determine incidence, as platinum-induced hearing loss is classically irreversible. All available audiograms were considered when determining onset and progression of hearing loss.

Descriptive statistics were calculated for demographic data and cumulative carboplatin dose at the time of last audiogram. Fisher’s exact test was performed to analyse the effect of sex (two-tailed) and exposure to individual risk factors (one-tailed) on incidence of clinically significant hearing loss (grade ≥2). Logistic regression was performed to analyse the effect of age at first exposure to carboplatin, cumulative carboplatin, and number of doses of carboplatin received, on grade ≥2 hearing loss. Given the small number of events, regression analyses were limited to univariate models. Receiver-Operating-Characteristic (ROC) curves were produced for these variables. (**Fig. 1**) The maximum Youden J value for each ROC curve was determined to find optimal cut-off points for these variables, and Fisher’s exact test was used to compare hearing outcomes in patients above and below these cut-offs. GraphPad Prism was used for all statistical analyses. P-values <0.05 were considered significant.

## Results

200 patients received carboplatin and had post-treatment audiological assessments available for review. 137/200 patients had pre-treatment audiograms available, all of which indicated normal hearing. The remaining 63 had no previously identified documented concerns for hearing loss. Clinical characteristics are described in **Table 2**. 104/200 (52%) patients were female. 101 patients (50.5%) had LGGs, 34 (17.0%) had retinoblastoma, 18 (9.0%) had intracranial germ cell tumours, 20 (10.0%) had extracranial germ cell tumours, and 27 (13.5%) had low- or intermediate-risk neuroblastoma. The median age at diagnosis was 35 months (range 0 days –17.0 years), and median age at first dose of carboplatin was 41 months (range 0 days – 17.0 years).

**Table 2:** Patient Demographics and Characteristics.

| Table 2: Patient Demographics and Characteristics |  |  |  |  |
| --- | --- | --- | --- | --- |
|  |  | All Patients (n=200) | SIOP Grade 0–1 (n=191) | SIOP Grade 2–4 (n=9) |
| **Sex, n (%)** |  |  |  |  |
|  | Male | 96 (48.0) | 92 (48.2) | 4 (44.4) |
|  | Female | 104 (52.0) | 99 (51.8) | 5 (55.6) |
| **Disease Group, n (%)** |  |  |  |  |
|  | Low-Grade Glioma | 101 (50.5) | 98 (51.3) | 3 (33.3) |
|  | Retinoblastoma | 34 (17.0) | 32 (16.8) | 2 (22.2) |
|  | Intracranial GCT | 18 (9.0) | 17 (8.9) | 1 (11.1) |
|  | Extracranial GCT | 20 (10.0) | 18 (9.4) | 2 (22.2) |
|  | Neuroblastoma | 27 (13.5) | 26 (13.6) | 1 (11.1) |
| **Age, median (range)** |  |  |  |  |
|  | Age at Diagnosis in months | 35.0 (0.0 – 204.5) | 39.2 (0.0 – 204.5) | 10.5 (0.4 – 135.7) |
|  | Age at First Carboplatin in months | 41.0 (0.2 – 204.8) | 44.0 (0.2 – 204.8) | 11.1 (0.4 – 135.9) |
|  | Age at Last Audiogram in months | 112.6 (3.2 – 354.5) | 113.9 (3.2 – 354.5) | 105.7 (12.1 – 167.3) |
| **Auditory follow-up, median (range)** |  |  |  |  |
|  | Auditory follow-up in months | 19.4 (0.0 – 188.7) | 17.7 (0.0 – 188.7) | 32.4 (6.3 – 151.1) |
| **Cumulative Carboplatin at last audiogram, median (range)** |  |  |  |  |
|  | Cumulative Carboplatin in mg/m<sup>2</sup> | 3366 (375 – 13335) | 3350 (375 – 13335) | 4407 (392 – 6570) |
| **Risk Factor Exposure at Last Audiogram, n (%)** |  |  |  |  |
|  | Vincristine | 69 (34.5) | 67 (35.1) | 2 (22.2) |
|  | Cranial Radiotherapy | 32 (16.0) | 30 (15.7) | 2 (22.2) |
|  | CSF Shunt | 31 (15.5) | 29 (15.2) | 2 (22.2) |
|  | Aminoglycosides | 65 (32.5) | 60 (31.4) | 5 (55.6) |
|  | Loop Diuretics | 17 (8.5) | 15 (7.9) | 2 (22.2) |
|  | Aminoglycosides or Loop Diuretics | 69 (34.5) | 64 (33.5) | 5 (55.6) |

The median age of the “normal hearing” group (SIOP grade 0–1) at first dose of carboplatin was 44 months (range 5 days – 17.0 years), while that of the “hearing loss” group (SIOP grade ≥2) was 11.1 months (range 12 days – 11.3 years). The median cumulative carboplatin dose was 3366mg/m^2^ in the whole cohort, 3350mg/m^2^ in the normal hearing group, and 4407mg/m^2^ in the hearing loss group. 69 patients (34.5%) were exposed to vincristine, 32 (16.0%) received cranial radiotherapy, 31 (15.5%) received a CSF shunt, and 69 (34.5%) received ototoxic drugs, i.e. aminoglycosides or loop diuretics.

805 audiograms of 200 patients were analysed. Hearing loss outcomes are displayed in **Table 3** as per the SIOP ototoxicity criteria. Of 200 patients, 15 (7.5%) developed measurable bilateral changes in auditory thresholds. All 15 patients had symmetrical hearing loss bilaterally. Of these fifteen patients, nine (60.0%; 4.5% of the total cohort) developed clinically significant hearing loss (SIOP grade ≥2). Three of these patients had low-grade glioma, two had retinoblastoma, three had germ cell tumours (two extracranial, one intracranial), and one had neuroblastoma.

**Table 3:**
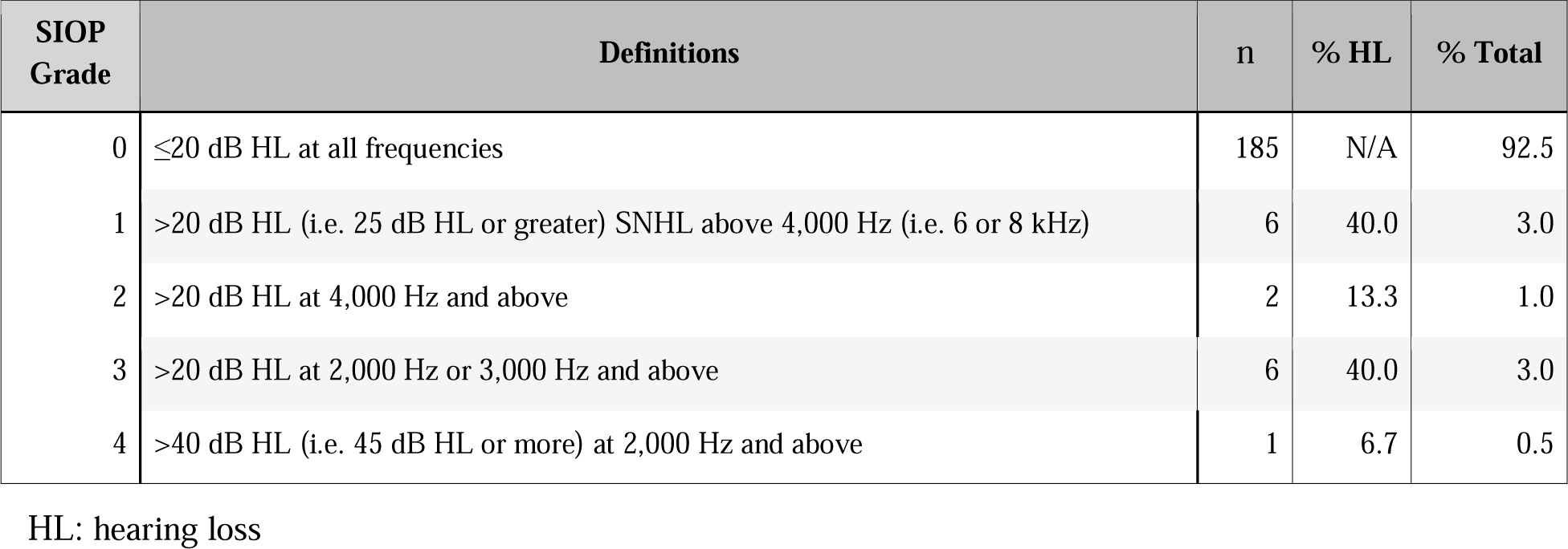
Hearing Loss Outcomes.

Progressive bilateral hearing loss was only observed in one patient, who progressed from grade 1 to grade 3 within six months. The median time from last carboplatin dose to first abnormal audiogram in these patients was 53 days (range 1 day – 12.2 years). Eleven additional patients (5.5%) developed unilateral auditory threshold changes, seven (3.5%) of which were classified as grade 1, three (1.5%) as grade 3, and one (0.5%) as grade 4.

The characteristics of patients who developed clinically significant bilateral hearing loss are displayed in **Table 4**. 8/9 patients who developed hearing loss were younger than 28 months (2.3 years) at time of first carboplatin exposure. Hearing aids were fitted in 8/9 of patients with hearing loss. Of these nine patients, all but one were exposed to other ototoxic treatments (**Table 4**). None of the identified binary risk factors (sex, exposure to vincristine, cranial radiotherapy, CSF shunting, or other ototoxic drugs) were significant predictors of clinically significant hearing loss (**Table 5**). Patients with “normal hearing” (SIOP grades 0-1) received auditory follow-up for a median of 17.7 months (range 0.0-188.7), while those with “hearing loss” (SIOP grades ≥2) received median auditory follow-up of 32.4 months (range 6.3-151.1).

**Table 4:** Characteristics of Patients with Hearing Loss at Last Audiogram (n=9).

|  | Characteristics |  |  |  | Risk Factors |  |  |  | Outcomes |  |
| --- | --- | --- | --- | --- | --- | --- | --- | --- | --- | --- |
|  | Sex | Disease Group | Age Group (years) | Cum. Carbo | Vincristine | Radiotherapy | CSF Shunt | Ototoxic Drugs | SIOP Grade | Hearing Aid |
| 1 | M | Low-Grade Glioma | 0–5 | 4658 | N | N | Y | N | 2 | N |
| 2 | M | Low-Grade Glioma | 0–5 | 4407 | N | Y | Y | N | 2 | Y |
| 3 | M | Low-Grade Glioma | 0–5 | 6570 | N | N | N | Y | 3 | Y |
| 4 | F | Retinoblastoma | 0–5 | 5889 | Y | N | N | N | 3 | Y |
| 5 | F | Retinoblastoma | 0–5 | 2717 | Y | N | N | Y | 3 | Y |
| 6 | F | Extracranial GCT | 0–5 | 6000 | N | N | N | Y | 3 | Y |
| 7 | F | Extracranial GCT | 0–5 | 4000 | N | N | N | N | 4 | Y |
| 8 | F | Intracranial GCT | 11–15 | 1802 | N | Y | N | Y | 3 | Y |
| 9 | M | Neuroblastoma | 0–5 | 392 | N | N | N | Y | 3 | Y |

**Table 5:** Effect of Risk Factors on Hearing Outcomes.

| Table 5: Effect of Risk Factors on Hearing Outcomes |  |  |
| --- | --- | --- |
|  | p-value | Odds Ratio (95% CI) |
| Sex | >0.9999 | 0.8609 (0.2579 to 2.982) |
| Vincristine | 0.3444 | 0.5288 (0.1088 to 2.491) |
| Cranial Radiotherapy | 0.3845 | 1.735 (0.3483 to 8.634) |
| CSF shunt | 0.4199 | 1.596 (0.3212 to 7.885) |
| Ototoxic Drugs | 0.1582 | 2.480 (0.7093 to 8.271) |
| Age | **0.0241** | 0.7888 (0.5620 to 0.9747) |
| Cumulative Carboplatin | 0.9429 | 1.000 (0.9997 to 1.000) |
| Number of Carboplatin Doses | 0.3888 | 1.062 (0.9217 to 1.207) |

Simple logistic regression showed that younger age at first dose of carboplatin was a significant predictor of hearing loss (p=0.0241, OR=0.7888) (**Table 5**); for each year of age, ototoxicity odds decreased by 21%. Cumulative carboplatin and number of carboplatin doses were not significant predictors of hearing loss. Corresponding ROC curves are displayed in **Supplementary Material S1**. Maximum Youden J values of these ROC curves showed optimal cut-off points for detecting hearing loss to be aged ≤28 months at first dose of carboplatin, >3984mg/m^2^ cumulative carboplatin, and <10 doses of carboplatin. Of these, the only statistically significant cut-off was age ≤28 months, as per Fisher’s exact test (p=0.0042, OR 12.37) (**Table 6**). 8/83 (9.6%) patients aged ≤28 months and 1/117 (0.85%) patients aged >28 months developed clinically significant hearing loss.

**Table 6:** Effect of Dichotomised Continuous Variables on Hearing Outcomes.

|  | p-value | Odds Ratio (95% CI) |
| --- | --- | --- |
| Age ≤ 28 months | **0.0042** | 12.37 (1.822 – 138.4) |
| Cumulative Carboplatin $>3984\text{mg/m}^2$ | 0.1245 | 0.3527 (0.09486 – 1.348) |
| No. of Carboplatin Doses $<10$ | 0.0787 | 5.287 (0.7813 – 59.41) |

## Discussion

Here we have demonstrated that clinically significant carboplatin-associated hearing loss is uncommon in children, affecting only nine patients (4.5%) in our cohort of 200. However, when present it may be severe and functionally limiting, with 8/9 (88.9%) of these children requiring hearing aids. Of these nine children, eight were younger than 28 months (2.3 years) at first exposure to carboplatin. Young age was the only significant predictor of hearing loss in this cohort, and ROC analysis identified 28 months (2.3 years) as the optimal exploratory cut-off.

The remaining one patient also received radiotherapy to their spine and skull, which may have contributed to their ototoxicity. We did not observe any hearing loss in patients above the age of 28 months who had not received cranial radiotherapy. This suggests that in our cohort, ototoxicity risk was concentrated in children younger than approximately 30 months at first carboplatin exposure.

Several biological mechanisms may plausibly contribute to the greater risk of ototoxicity observed in younger children. These include increased permeability of the blood–labyrinth barrier, potentially resulting in greater cochlear platinum exposure, and less mature antioxidant defences against reactive oxygen species (ROS), which are implicated in platinum-induced ototoxicity.^14,16^ Additionally, younger children are dosed according to renal function (creatinine-based eGFR) and target area-under-the-curve (AUC) approaches in some protocols, but age-related pharmacokinetic variability may lead to carboplatin overexposure.^17^ None of these mechanisms can readily account for the specific cut-off identified in our cohort.

In our cohort, a small number of patients (n=4; 2.0%) demonstrated clinically significant unilateral hearing loss (SIOP grade ≥2). When considering these patients, overall incidence of clinically significant hearing loss would be 6.5% (n=13). However, these patients were not included in our primary outcome analysis as unilateral hearing loss is not typical for platinum-induced ototoxicity and is more likely to reflect alternative etiologies such as middle ear pathology, intercurrent illness, or measurement variability. This approach is consistent with previous literature that considers platinum-induced hearing loss to be typically symmetrical and bilateral, but may under-report the true incidence of hearing loss when compared to studies using the worse ear.

Our results are most similar to those of Jehanne et al. (n=175), who recorded hearing loss in 4.6% of their patients.^18^ (**Table 1**) Qaddoumi et al. (n=60) and Soliman et al. (n=71) both demonstrated significantly higher incidences of hearing loss (16.7% and 33.8%, respectively).^4,19^ However, these studies only evaluated patients diagnosed with retinoblastoma, and this population tends to be younger than our cohort. Qaddoumi et al.’s cohort had a median age at diagnosis of 8.6 months, whereas our median age at diagnosis was 35 months. This difference, alongside our finding that young age is a significant risk factor, may explain why their reported incidence of hearing loss is higher than in our cohort. Only 2/34 (5.9%) of our cohort with retinoblastoma developed hearing loss, but we were limited by a smaller sample size of patients with retinoblastoma. Unlike Musial-Bright et al. and Edward et al., we did not observe a relationship between cumulative dose of carboplatin and hearing loss.^5,20^ However, our patient population did not receive high dose carboplatin (e.g. for myeloablative conditioning prior to stem cell transplant), which may present a different risk profile for ototoxicity and warrants further investigation.

Accurate comparison between studies is limited due to different definitions of hearing loss. Qaddoumi et al. defined hearing loss as grade ≥1 on any one of the NCI-CTCAE v3, Brock, or CCG criteria, while Soliman et al. defined hearing loss as grade ≥1 on of the SIOP, Brock, Chang, ASHA, or NCI-CTCAE criteria. Our study defined “clinically significant hearing loss” as grade ≥2 on the SIOP ototoxicity scale, which has demonstrated increased sensitivity compared to other criteria in identifying hearing loss. ^21^ Grade 2 was used as the cutoff as changes in auditory thresholds at frequencies above 4kHz (i.e. SIOP grade 1) do not classically correlate with functional loss in hearing, particularly as human speech consonants occur predominantly within the 0.5-4kHz ranges.^21^ A comparison of different grading criteria can be found in **Supplementary Material S2**. Furthermore, while Musial-Bright et al. used the better ear to classify hearing loss (in line with our methodology), Soliman et al. used the worse ear, and in the other three noted articles this was not specified. Our study used the better hear to reflect functional hearing, and exclude isolated unilateral hearing losses which are not typical of platinum-induced ototoxicity. These discrepancies may influence reported hearing outcomes as use of the worse ear would also include unilateral hearing losses, which are not typical for platinum-induced hearing loss.

Progressive and late-onset hearing loss has been observed in patients receiving cisplatin but infrequently described in association with carboplatin. Peleva et al. reported progression of hearing loss in 48% of children receiving platinum compounds (cisplatin, carboplatin, or both), as assessed by the ASHA criteria.^23^ Lambert et al. noted that one child in their cohort of 116 had progressive loss, worsening from unilateral to bilateral after one dose of carboplatin.^24^ In our cohort, one patient progressed from grade 1 to grade 3 hearing loss within six months; during this period, the patient received two doses of carboplatin and also received 14 doses of gentamicin which may have contributed to their ototoxicity. The median time from last carboplatin dose to first abnormal audiogram (SIOP grade ≥2 bilaterally) was 53 days (range 1 day – 12.2 years). However, this is not an accurate proxy for true onset to ototoxicity as audiograms occurred at discrete time points, and the true onset of ototoxicity may have occurred between audiograms. Furthermore, our hearing outcomes were censored to date of last audiogram, and our “normal hearing” group received shorter auditory follow-up overall than our “hearing loss” group (17.7 months vs 32.4 months). This likely reflects ongoing audiological monitoring in patients after hearing loss had been identified. However, patients with initially normal hearing may have developed late-onset hearing loss not captured by our study, particularly in the absence of routine audiological surveillance.

The main strengths of our study are our large sample size (n=200) and our use of the SIOP ototoxicity criteria to grade audiograms. This is the largest study to date of carboplatin-associated hearing loss in cisplatin-naïve children. Furthermore, many comparable studies predate the SIOP criteria, limiting its use in the existing literature. Lastly, we also evaluated co-exposures that have been recently implicated in platinum-associated ototoxicity (vincristine, CSF shunts) but have not previously been investigated in a carboplatin-specific context.

As in prior studies, concomitant ototoxic exposures were a key limitation of our study. Although we did not observe a significant additive effect of vincristine, cranial radiotherapy, CSF shunts, or ototoxic drugs, these co-exposures may have confounded our hearing outcomes. Only one patient with hearing loss had not been exposed to any of these treatments at the time of their last audiogram. Also, as baseline audiology was not available for 63/200 patients, five of whom developed hearing loss, we cannot be confident that their hearing loss was attributable to carboplatin. Thus, the true incidence of carboplatin-induced hearing loss is likely even lower than what we are reporting. Determining onset and progression of hearing loss was also difficult in our cohort, as hearing was assessed at discrete, non-standardised timepoints. This limitation can be addressed in the future through a prospective study design.

Despite being uncommon, carboplatin-induced hearing loss can still have significant impacts on patients’ development and quality of life. Hearing surveillance is generally recommended for children receiving platinum chemotherapy, with monitoring intensity tailored to regimen and risk but this may not be feasible, especially for resource-limited centres. Our results have shown that younger children, particularly those aged less than approximately 30 months (2.5 years) at first carboplatin exposure, are significantly more likely to develop hearing loss. We therefore recommend stringent formal audiological monitoring for younger children receiving carboplatin, as well as continued surveillance for any child with identified auditory threshold changes. Less resource-intensive ways of monitoring hearing, such as subjective questionnaires and screening tools, may be considered for older children, in accordance with local survivorship guidelines and clinical context. Further validation of our results is recommended through prospective studies with a larger sample size and/or with a smaller proportion of patients exposed to confounding treatments. Additionally, investigation of incidence of ototoxicity in children receiving high-dose carboplatin is recommended.

In summary, our study has shown that clinically significant carboplatin-associated hearing loss is uncommon in children (incidence 4.5%). However, we may be overestimating incidence as our study was limited by missing baseline audiological assessments and concomitant ototoxic exposures. Similarly, use of the better ear and classification of hearing loss as SIOP grade ≥2 in may cause us to under-report true ototoxicity potential, despite reflecting functional hearing outcomes. Younger age at first exposure to carboplatin was associated with hearing loss (OR 0.7888, p=0.0241). Children aged ≤28 months were identified as at greatest risk of hearing loss (OR 12.37, p=0.0042). We can use this finding to better counsel parents regarding risk of hearing loss, while also optimising resource allocation and targeting surveillance towards younger children, particularly those who receive carboplatin at age ≤30 months.

## Data Availability

All data produced in the present study are available upon reasonable request to the authors.

## Acknowledgements

We extend our gratitude to the individuals and institution at the University of Auckland, whose support has allowed the completion of this work. DDE was funded by an Establishment grant from the Royal Children’s Hospital Foundation (2019-1193).

## Abbreviations

AUC: Area under the curve
CCG: Children’s Cancer Group
CSF: Cerebrospinal fluid
GCT: Germ Cell Tumour
LGG: Low-grade glioma
HL: Hearing loss
OR: Odds Ratio
ROC: Receiver Operating Characteristic
SIOP: International Society of Paediatric Oncology
STS: Sodium thiosulfate

## References

1. Ruggiero A, Trombatore G, Triarico S, Arena R, Ferrara P, Scalzone M, Pierri F, Riccardi R. Platinum compounds in children with cancer: toxicity and clinical management. Anti-cancer drugs. 2013 Nov 1;24(10):1007–19.

2. Knight KR, Kraemer DF, Neuwelt EA. Ototoxicity in children receiving platinum chemotherapy: underestimating a commonly occurring toxicity that may influence academic and social development. Journal of Clinical Oncology. 2005 Dec 1;23(34):8588–96.

3. Batra A, Thakar A, Bakhshi S. Ototoxicity in retinoblastoma survivors treated with carboplatin based chemotherapy: a cross sectional study of 116 patients. Pediatric blood & cancer. 2015 Nov;62(11):2060-.

4. Qaddoumi I, Bass JK, Wu J, Billups CA, Wozniak AW, Merchant TE, Haik BG, Wilson MW, Rodriguez-Galindo C. Carboplatin-associated ototoxicity in children with retinoblastoma. Journal of Clinical Oncology. 2012 Apr 1;30(10):1034–41.

5. Musial-Bright L, Fengler R, Henze G, Hernáiz Driever P. Carboplatin and ototoxicity: hearing loss rates among survivors of childhood medulloblastoma. Child’s Nervous System. 2011 Mar;27:407–13.

6. Fetoni AR, Ruggiero A, Lucidi D, De Corso E, Sergi B, Conti G, Paludetti G. Audiological monitoring in children treated with platinum chemotherapy. Audiology and Neurotology. 2016 Jun 11;21(4):203–11.

7. Murphy B, Jackson A, Bass JK, Tsang DS, Ronckers CM, Kremer L, Baliga S, Olch A, Zureick AH, Jee KW, Constine LS. Modeling the risk of hearing loss from radiation therapy in childhood cancer survivors: A PENTEC comprehensive review. International Journal of Radiation Oncology* Biology* Physics. 2023 Oct 19.

8. Moke DJ, Luo C, Millstein J, Knight KR, Rassekh SR, Brooks B, Ross CJ, Wright M, Mena V, Rushing T, Esbenshade AJ. Prevalence and risk factors for cisplatin-induced hearing loss in children, adolescents, and young adults: a multi-institutional North American cohort study. Lancet Child Adolesc Health. 2021; 5 (4): 274–83.

9. Strebel S, Mader L, Jörger P, Waespe N, Uhlmann S, von der Weid N, Ansari M, Kuehni CE. Hearing loss after exposure to vincristine and platinum-based chemotherapy among childhood cancer survivors. EJC Paediatric Oncology. 2023 Jan 1;1:100017.

10. Meijer AJ, Li KH, Brooks B, Clemens E, Ross CJ, Rassekh SR, Hoetink AE, Van Grotel M, van den Heuvel-Eibrink MM, Carleton BC. The cumulative incidence of cisplatin-induced hearing loss in young children is higher and develops at an early stage during therapy compared to older children, based on 2,052 audiological assessments. Childhood cancer related hearing loss and tinnitus. 2021 Nov 1:41.

11. Guillaume DJ, Knight K, Marquez C, Kraemer DF, Bardo DM, Neuwelt EA. Cerebrospinal fluid shunting and hearing loss in patients treated for medulloblastoma. Journal of Neurosurgery: Pediatrics. 2012 Apr 1;9(4):421–7.

12. Schacht J, Talaska AE, Rybak LP. Cisplatin and aminoglycoside antibiotics: hearing loss and its prevention. The Anatomical Record: Advances in Integrative Anatomy and Evolutionary Biology. 2012 Nov;295(11):1837–50.

13. Ding D, Liu H, Qi W, Jiang H, Li Y, Wu X, Sun H, Gross K, Salvi R. Ototoxic effects and mechanisms of loop diuretics. Journal of otology. 2016 Dec 1;11(4):145–56.

14. Brock PR, Knight KR, Freyer DR, Campbell KC, Steyger PS, Blakley BW, Rassekh SR, Chang KW, Fligor BJ, Rajput K, Sullivan M. Platinum-induced ototoxicity in children: a consensus review on mechanisms, predisposition, and protection, including a new International Society of Pediatric Oncology Boston ototoxicity scale. Journal of Clinical Oncology. 2012 Jul 1;30(19):2408–17.

15. Clemens E, Brooks B, De Vries AC, Van Grotel M, Van Den Heuvel-Eibrink MM, Carleton B. A comparison of the Muenster, SIOP Boston, Brock, Chang and CTCAEv4. 03 ototoxicity grading scales applied to 3,799 audiograms of childhood cancer patients treated with platinum-based chemotherapy. PLoS One. 2019 Feb 14;14(2):e0210646.

16. Romano A, Capozza MA, Mastrangelo S, Maurizi P, Triarico S, Rolesi R, Attinà G, Fetoni AR, Ruggiero A. Assessment and management of platinum-related ototoxicity in children treated for cancer. Cancers. 2020 May 17;12(5):1266.

17. van de Velde ME, den Bakker E, Blufpand HN, Kaspers GL, Abbink FC, Kors AW, Wilhelm AJ, Honeywell RJ, Peters GJ, Stoffel-Wagner B, Buffart LM. Carboplatin dosing in children using estimated glomerular filtration rate: equation matters. Cancers. 2021 Nov 26;13(23):5963.

18. Jehanne M, Lumbroso Le Rouic L, Savignoni A, Aerts I, Mercier G, Bours D, Desjardins L, Doz F. Analysis of ototoxicity in young children receiving carboplatin in the context of conservative management of unilateral or bilateral retinoblastoma. Pediatric blood & cancer. 2009 May;52(5):637–43.

19. Soliman SE, D’Silva CN, Dimaras H, Dzneladze I, Chan H, Gallie BL. Clinical and genetic associations for carboplatin related ototoxicity in children treated for retinoblastoma: a retrospective noncomparative single institute experience. Pediatric Blood & Cancer. 2018 May;65(5):e26931.

20. Edward ED, Rosdiana N, Farhat F, Siregar OR, Lubis B. Prevalence and risk factors of hearing loss in children with solid tumors treated with platinum-based chemotherapy. Paediatrica Indonesiana. 2015 Jun 30;55(3):121–5.

21. Knight KR, Chen L, Freyer D, Aplenc R, Bancroft M, Bliss B, Dang H, Gillmeister B, Hendershot E, Kraemer DF, Lindenfeld L. Group-wide, prospective study of ototoxicity assessment in children receiving cisplatin chemotherapy (ACCL05C1): a report from the Children’s Oncology Group. Journal of clinical oncology. 2017 Feb 1;35(4):440–5.

22. Walker JJ, Cleveland LM, Davis JL, Seales JS. Audiometry screening and interpretation. Am Fam Physician. 2013;87(1):41–47.

23. Peleva E, Emami N, Alzahrani M, Bezdjian A, Gurberg J, Carret AS, Daniel SJ. Incidence of platinum induced ototoxicity in pediatric patients in Quebec. Pediatric blood & cancer. 2014 Nov;61(11):2012–7.

24. Lambert MP, Shields C, Meadows AT. A retrospective review of hearing in children with retinoblastoma treated with carboplatin based chemotherapy. Pediatric blood & cancer. 2008 Feb;50(2):223–6.

