## Supplementary Material S1 for "Incidence and Severity of Carboplatin-Associated Hearing Loss in Children with Cancer Assessed by the SIOP 2012 Ototoxicity Criteria"

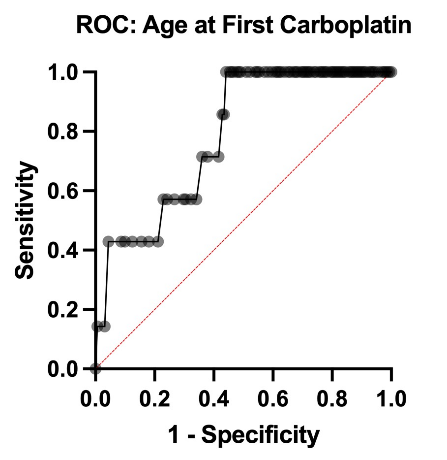

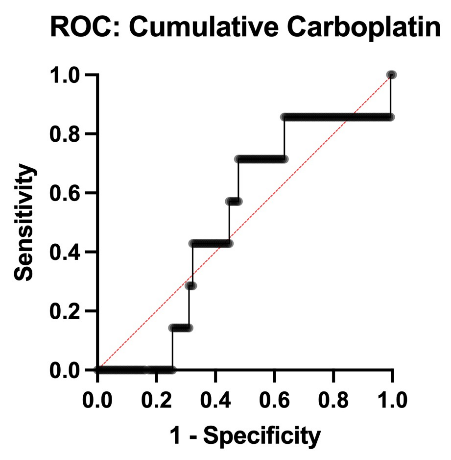

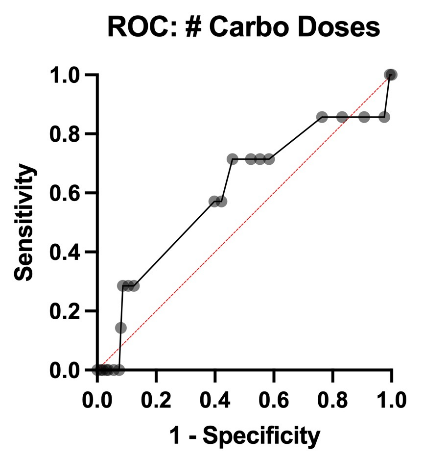

Supplementary Figure 1: ROC Curves for Age at First Exposure, Cumulative Carboplatin, and Number of Doses

A

C

B
