## Supplementary Material S2 for "Incidence and Severity of Carboplatin-Associated Hearing Loss in Children with Cancer Assessed by the SIOP 2012 Ototoxicity Criteria"

| Supplementary Table 1: Ototoxicity Grading Criteria | | | | | | |
| --- | --- | --- | --- | --- | --- | --- |
|  | Brock | ASHA | NCI-CTCAE v6.0* | Chang | SIOP Ototoxicity Scale | Children’s Cancer Group |
| 0 | <40dB at all frequencies | Binary system, HL if:   1. 20 dB decrease at any one test frequency, or 2. 10 dB decrease at any two adjacent test frequencies, or 3. Loss of response at three consecutive test frequencies where responses were previously obtained |  | ≤20dB at 1, 2, and 4 kHz | ≤20dB at all frequencies | No hearing loss |
| 1 | ≥40dB at 8000Hz |  | >20dB at 6kHz or 8kHz in at least one ear | 1a: ≥40dB at any frequency 6-12 kHz  1b: >20 and <40dB at 4kHz | >20dB SNHL above 4000Hz (i.e. 6 or 8kHz) | ≥40dB at 6000 and/or 8000 Hz |
| 2 | ≥40dB at 4000Hz and above |  | >20dB at 4kHz in at least one ear; mild/moderate impact on age-appropriate normal daily activity | 2a: ≥40dB at 4kHz and above  2b: >20 and <40dB at any frequency below 4kHz | >20dB SNHL at 4000Hz and above | >25dB at 3000 and/or 4000 Hz |
| 3 | ≥40dB at 2000Hz and above |  | Hearing loss sufficient to indicate therapeutic intervention (e.g. aids) or >20dB at 2kHz or 3kHz in at least one ear; severe impact on age-appropriate normal daily activity | ≥40 dB at 2 or 3 kHz and above | >20dB SNHL at 2000Hz or 3000Hz and above | >25dB at 2000 Hz |
| 4 | ≥40dB at 1000Hz and above |  | Audiologic indication for cochlear implant; >40dB HL with SNHL at 2kHz and above | ≥40 dB at 1 kHz and above | >40dB SNHL at 2000Hz and above | ≥40dB at 2000 Hz |

* Latest iteration of NCI-CTCAE criteria (v6.0); previous versions may have used in earlier literature.
